# Testosterone Effects on Short-Term Physical, Hormonal, and Neurodevelopmental Outcomes in Infants with 47,XXY/Klinefelter Syndrome: The TESTO Randomized Controlled Trial

**DOI:** 10.1101/2024.12.09.24318726

**Authors:** Shanlee Davis, Susan Howell, Jennifer Janusz, Najiba Lahlou, Regina Reynolds, Talia Thompson, Karli Swenson, Rebecca Wilson, Judith Ross, Philip Zeitler, Nicole Tartaglia

## Abstract

**Context:** 47,XXY/Klinefelter syndrome (XXY) is associated with impaired testicular function and differences in physical growth, metabolism, and neurodevelopment. Clinical features of XXY may be attributable to inadequate testosterone during the mini-puberty period of infancy.

**Objective:** We tested the hypothesis that exogenous testosterone treatment positively effects short-term physical, hormonal, and neurodevelopmental outcomes in infants with XXY.

**Design:** Double-blind randomized controlled trial, 2017-2021

**Setting:** US tertiary care pediatric hospital

**Patients:** Infants 30-90 days of age with prenatally identified, non-mosaic 47,XXY (n=71).

**Intervention:** Testosterone cypionate 25mg intramuscular injections every 4 weeks for 3 doses

**Main outcome measures:** The *a priori* primary outcomes were change in percent fat mass (%FM) z-scores and change in the total composite percentile on Alberta Infant Motor Scales (AIMS) assessment from baseline to 12 weeks.

**Results:** The between group difference in change in %FM z-scores was -0.57 [95% CI -1.1, - 0.06], p=0.03), secondary to greater increases in lean mass in the testosterone-treated group (1.5±0.4 kg vs 1.2±0.4, p=0.001). Testosterone suppressed gonadotropins and inhibin B (p<0.001 for all). In contrast, there were no significant group differences in short term motor, cognitive, or language outcomes (p>0.15 for all).

**Conclusions:** In this double-blind randomized controlled trial in infants with XXY, testosterone injections resulted in physical effects attributable to systemic androgen exposure; however, there was no impact on neurodevelopmental outcomes and the hypothalamic-pituitary-gonadal axis was suppressed. These results do not support routine testosterone treatment in infants with XXY, however long term follow up on physical health, neurodevelopment and testicular function is needed.

## INTRODUCTION

The widespread adoption of noninvasive prenatal screening (NIPS) via cell-free DNA brought with it a substantial increase in the number of infants recognized to have sex chromosome aneuploidies.(1) The most common sex chromosome aneuploidy, 47,XXY or Klinefelter syndrome, is estimated to affect in 1 in 600 males.(2) Most infants born with XXY have no overt signs or symptoms, although some studies report subtle, nonspecific features in comparison to males without XXY including smaller birth size, reduced penile growth in the first year of life, mild hypotonia, and a more passive temperament.(3) As adolescents and adults, individuals with XXY typically have testicular dysfunction, which manifests as microorchidism, hypogonadotropic hypogonadism, and infertility.(4) Neurocognitive and cardiometabolic manifestations are also well-described as prominent features of the phenotype contributing to increased morbidity throughout childhood and adulthood, as well as increased mortality.(2,5–11) While testosterone deficiency can contribute to neurocognitive deficits and poor cardiometabolic health in men, the causal role in XXY is unclear as post-pubertal testosterone treatment fails to normalize these findings. With these observations, in addition to more individuals being diagnosed earlier in life, there is a growing interest in prevention and/or early intervention opportunities to decrease morbidity in XXY.

In the first months of life, the hypothalamic-pituitary-gonadal (HPG) axis is transiently active, driving testicular testosterone production in males.(12) Although the specific purpose of this mini-puberty period of infancy is unclear, it is hypothesized to be a critical window in development with an essential role in sexual differentiation of multiple tissues and programming for future sex-specific cellular processes.(3,13–21) Several human studies have reported associations between infant testosterone production and sex differences in linear growth, lean mass accumulation, brain lateralization, language organization, and gender identity.(17,22,23) Mouse models manipulating sex steroid exposure have bolstered these findings, illustrating that early postnatal testosterone exposure stimulates systemic epigenetic changes resulting in permanent impacts on neurocognition and energy metabolism.(14,16) Studies in infants with XXY confirm that the mini-puberty testosterone surge does occur, however, systemic testosterone concentrations are lower than average.(24–27) Furthermore, retrospective reports of a clinical cohort of boys with XXY treated with testosterone in infancy describe higher cognitive, language, motor, and social communication abilities in childhood compared to boys without a history of testosterone treatment.(19,28) Previously, we reported differences in adiposity between infants with XXY who did and did not receive testosterone.(29) However, there has not been a rigorously conducted study to inform the potential benefits or risks of infant testosterone treatment in XXY.

The aim of this double-blind, placebo-controlled, randomized prospective clinical trial was to test the hypothesis that exogenous testosterone treatment during the mini-puberty period of infancy has positive effects on short-term physical, hormonal, and neurodevelopmental outcomes in boys with XXY. Additional aims were to inform the side effect profile of treatment, determine if any observed immediate effects are sustained, and explore whether the timing (age) of intervention matters. This study was designed to inform best clinical practice recommendations for the many infant boys now prenatally identified to have an additional X chromosome.

## METHODS

### Overall Study Design

This was a randomized controlled trial assessing the efficacy and safety of testosterone injections in infants with XXY. Infants with prenatally identified, non-mosaic 47,XXY received either testosterone or placebo injections for three months followed by three months of cross-over intervention. Outcomes were assessed by blinded investigators at baseline, after the first treatment period (12 weeks), and after the cross-over period (24 weeks).

### Setting, Recruitment, and Participants

The study took place at Children’s Hospital Colorado (CHCO) in the outpatient Pediatric Clinical Translational Research Center (CTRC). Participants were recruited through the interdisciplinary eXtraordinarY Kids Clinic at CHCO, local genetics and obstetrics offices, local and national advertisements, XXY social media groups, family support groups, and national pediatric endocrinology mailing lists. Infants between 31-90 days of age were eligible if they were prenatally identified and subsequently confirmed via chorionic villous sampling, amniocentesis, or postnatal blood/tissue to have a karyotype of 47,XXY. Exclusion criteria included >20% mosaicism for typical 46,XY cell line, gestational age <36 weeks, birth weight <2.5% or >97.5% for age, use of medications known to impact body composition (e.g. insulin, growth hormone), allergy to any of the components in testosterone cypionate, history of thrombosis in self or first-degree relative, and exposure to androgen therapy outside of the study protocol. The study was approved by the local institutional review board (COMIRB 17-1317), registered on ClinicalTrials.gov (NCT03325647), and the protocol was filed with the US Food and Drug Administration (Investigative New Drug file #124260). The parents of every participant provided written informed consent prior to any study procedures. An independent data safety and monitoring board (DSMB) provided additional study oversight.

### Randomization

Following enrollment, participants were assigned to one of two groups through block randomization in blocks of 20 with a 1:1 allocation scheme determined from an automated list generated at the beginning of the study. To ensure allocation concealment, CHCO Investigational Drug Services generated and held the randomization list and dispensed the study drug. All investigators and parents remained blinded to both the allocation as well as the intervention until all study visits were completed and the data were locked for analysis. No interim analyses for efficacy were conducted.

### Intervention

Participants received a total of six 0.125 ml injections in the vastus muscle during the six months study period. Participants randomized to Group A were given one injection of 25 mg testosterone cypionate (200 mg/mL concentration) every 28 days for a total of three injections, follow by placebo (saline) injection once every 28 days for a total of three injections during study weeks 12-24. Group B received placebo injections every 28 days for the first three months of the study, followed by three testosterone injections. This testosterone regimen was chosen based on its efficacy and safety in treating boys with micropenis.(30)

The first injection was administered by a nurse at the end of the initial study visit. If it was not feasible to return for monthly injections, parents were trained to give subsequent injections themselves and were given a dosing calendar and log. A prepared syringe with study drug was mailed to the participant for each dose. An automated reminder email was sent to the parents 24 hours before each injection was due, and an electronic survey was sent one week following the injection to document administration details and any side effects.

### Study Visits and Timeline

In-person study visits occurred at enrollment (baseline), 12 weeks (+/- 2 weeks), and 24 weeks (+/- 2 weeks). If an in-person study visit was not possible (e.g. secondary to the COVID-19 pandemic), the study visit was conducted via video conference. Enrollment commenced in November 2017 and final study visits occurred in May 2021.

### Study Assessments

Prenatal, birth, medical, developmental, and feeding histories were obtained from parents at the initial study visit. Any updates, including new medications and any perceived adverse events, were collected at subsequent visits. In addition, parents subjectively rated their infant’s temperament as “easy going”, “average”, or “difficult” at each visit. Physical exams were conducted by a board-certified pediatrician blinded to participant treatment status. Weight was measured to the nearest 0.01 kilogram. Stretched penile length, head circumference, arm span, waist circumference, and total body length were measured to the nearest 0.1 centimeter. Weight, length, head circumference and weight-for-length age-and sex-specific z-scores were calculated from World Health Organization (WHO) norms. Penile length z-scores were calculated from published norms.(31) Parent(s) completed a sociodemographic survey that included self-reported race, ethnicity, education level, occupation, and family income; answers were used to calculate the Hollingshead index as a measure of socioeconomic status (SES).

Body composition was assessed using air displacement plethysmography (PEAPOD) for infants up to 10 kg.(32) Using the measured body mass and total volume, fat mass (FM) and fat free mass (FFM) were estimated to the nearest gram, and %FM was then calculated by dividing the FM by the total body mass. Two independent PEADPOD measurements were obtained, with a third measurement if the first two %FM differed by >3%. Due to the vast differences in body composition expected in early infancy, sex- and age-specific %FM z-scores were calculated from published norms.(33)

Standardized neurodevelopmental assessments were administered by trained psychometrists, child psychologists, or pediatricians blinded to participant treatment status. The Alberta Infant Motor Scale (AIMS) is a measure of gross motor maturation designed to evaluate gross motor development over the first year of life in the prone, supine, sitting, and standing positions, with total scores ranging from 0 to 55 which is then converted to an age-normed percentile (0–100).(34,35) As the AIMS is observational, it is feasible to conduct in person or via video conference. The Peabody Developmental Motor Scales - Version 2 (PDMS-2) includes six subtests that assess gross and fine motor scales from birth to five years of age, yielding age-normed quotient scores (mean 100, SD 15) for total, fine, and gross-motor abilities and scaled scores (mean 10, SD 3) for each subdomain.(36) The Bayley Scales of Infant and Toddler Development, 3^rd^ edition (Bayley-3) is a well-validated standardized assessment spanning three domains: cognitive, language (expressive and receptive), and motor (fine and gross).(37) Raw scores were converted to age-normed composite scores (mean 100, SD 15) and subdomain scaled scores (mean 10, SD 3). In addition, Growth Score Values (GSV) based on raw scores without age comparison were used to assess change in development relative to each individual.(38) The PDMS-2 and Bayley-3 require in-person administration.

Parents completed the Adaptive Behavior Assessment System Third Edition (ABAS-3), a validated parent-report measure capturing adaptive skills across the life span. The ABAS-3 yields scaled scores in seven subdomains that inform three domain standard scores (mean 100, SD 15) and an overall Generalized Adaptive Composite (GAC). Parents also completed an electronic survey one week after every injection to document administration details, any deviations or problems with administration, and treatment emergent adverse events (from a provided list as well as open-ended). The severity and causality for all adverse events was determined by the study team; adverse events were regularly assessed by the DSMB. Infants had a venous blood draw in the morning following three hours of fasting at each study visit. Samples were collected in a serum separate tube, processed, and stored at -80 degrees Celsius until batch analysis without any freeze-thaw cycles.

### Laboratory Methods

Detailed methods for measurement of serum hormones can be found in the Supplement. In brief, total testosterone (TT) was measured using liquid chromatography-tandem mass spectrometry (LC-MS/MS) with a detection limit of 0.02 nmol/L; intra-assay coefficient of variation (CV) were <5% and inter-assay CVs were <8% throughout the range of observed values. Luteinizing hormone (LH) and follicle stimulating hormone (FSH) were measured by means of a sensitive electrochemiluminescent immunoassays with a quantification range 0.1-200 mIU/mL and cross-reactivity for similar hormones <0.1%. Inhibin B (INHB) and anti-mullerian hormone (AMH) were measured by solid-phase sandwich assays. The lower limit of detection for INHB was 4.6 pg/ml with a quantification range up to 1100 pg/ml. The lower limit of detection for AMH was 0.7 pmol/L with a quantification range up to 114 pmol/L. Extensive internal and external quality control measures were used for all assays (see Supplement).

### Statistical Analyses

Data were examined for outliers, missing values and normality by visual inspection and applicable tests; following these tests no outliers were removed, and the minimal missing data were treated as missing without imputation. Summary baseline data are reported as mean with standard deviation (SD), median with interquartile range (IQR), or number and proportion depending on data type. Baseline variables were compared between randomized groups to evaluate whether there were any intrinsic differences that would need to be accounted for in analyses.

The *a priori* primary outcomes were change in percent fat mass (%FM) z-scores from baseline to 12 weeks and change in the total composite percentile on the AIMS from baseline to 12 weeks.

Secondary and exploratory outcomes included change scores from both baseline to 12 weeks and 12-24 weeks for other body composition variables, anthropometric measurements, and standard scores on the PDMS, Bayley 3, and ABAS 3; serum hormone concentrations; and number and type of adverse events. Change scores were chosen as we predicted the intraindividual correlation to be high for most of our outcomes of interest, therefore allowing for a more precise estimate as well as clinical applicability in determining the effect of testosterone treatment.

All outcomes were assessed between groups with two-tailed t-tests or Wilcoxon tests along with computed difference in means (or medians) and 95% confidence intervals (CI), with significance set at 5%, adhering to an intention-to-treat analysis. An initial sample size of 27 per group was determined based on the ability to detect a %FM z-score difference of 0.5 between groups with 80% power; due to the COVID-19 pandemic, we overenrolled beyond this goal to account for anticipated missing longitudinal PEAPOD data. Following the primary unadjusted outcome analysis, linear regression models were performed with treatment group, baseline measure, and potential covariates (e.g. age of enrollment, duration between measures, feeding source, SES, baseline testosterone concentration) for all outcomes from both the 12- and 24-week visits as secondary/exploratory endpoints. Adverse events were summed and risk ratios with 95% confidence intervals calculated based on whether the participant was receiving testosterone or placebo at the time of the adverse event. All analyses were conducted in R v.4.3.2.

## RESULTS

The CONSORT diagram for the seventy-two enrolled participants in the TESTO trial is shown in Figure 1. The observed attrition was only 2.8%, however, not all outcome assessments could be completed due to the disruption caused by the COVID19 pandemic. Demographics and baseline assessments were similar between the randomized groups (Table 1). Infants were low-average size for gestational age and enrolled at a mean age of two months. One third of participants endorsed Hispanic ethnicity and/or non-White race, however the majority of the sample were in households making >$100,000 per year and had an average Hollingshead socioeconomic index that reflected a high level of parental education and occupational status. All but one had a prenatal cell free DNA screening positive for 47,XXY. Approximately half elected prenatal diagnostic testing (chorionic villous sampling or amniocentesis) while the other half confirmed the 47,XXY diagnosis postnatally. The one infant that was not identified by cell free DNA screening was diagnosed with elective amniocentesis for advanced maternal age.

**Figure 1.**
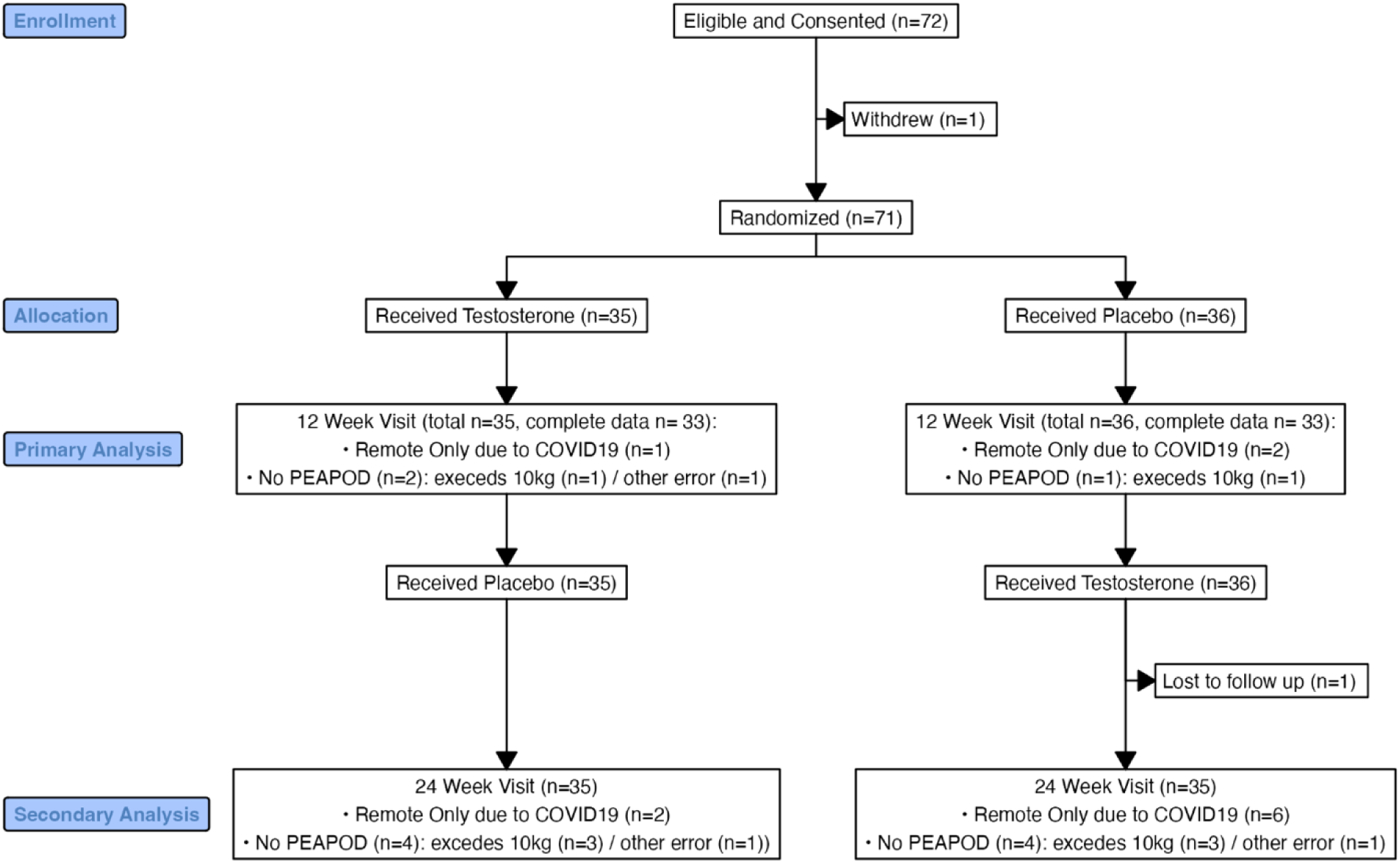
CONSORT diagram detailing participant allocation and completion of primary outcome measures for the TESTO trial.

**Table 1.** Baseline demographics and characteristics of the sample stratified by randomization group.

|  | All<br>(n=71) | Group A<br>(n=35) | Group B<br>(n=36) | p-value |
| --- | --- | --- | --- | --- |
| *Demographic Variables* |  |  |  |  |
| Race |  |  |  | 0.508 |
| White | 63 (88.7%) | 30 (85.7%) | 33 (91.7%) |  |
| Asian | 3 (4.2%) | 1 (2.9%) | 2 (5.6%) |  |
| Black | 4 (5.6%) | 3 (8.6%) | 1 (2.8%) |  |
| Native American | 1 (1.4%) | 1 (2.9%) | 0 |  |
| Ethnicity |  |  |  | 0.396 |
| Hispanic/Latino | 16 (22.5%) | 6 (17.1%) | 10 (27.8%) |  |
| Not Hispanic/Latino | 55 (77.5%) | 29 (82.9%) | 26 (72.2%) |  |
| Hollingshead SES Index | 55.5 [49, 61] | 55.5 [52, 61] | 53.8 [41, 61] | 0.245 |
| Household annual income |  |  |  | 0.217 |
| <\$50,000 | 2 (2.9%) | 0 | 2 (5.7%) | |
| \$50,000-75,000 | 9 (13.2%) | 4 (12.1%) | 5 (14.3%) | |
| \$75,000-100,000 | 10 (14.7%) | 4 (12.1%) | 6 (17.1%) | |
| \$100,000-150,000 | 17 (25.0%) | 11 (33.3%) | 6 (17.1%) | |
| \$150,000-250,000 | 18 (26.5%) | 6 (18.2%) | 12 (34.3%) | |
| >\$250,000 | 12 (17.6%) | 8 (24.2%) | 4 (11.4%) | |
| *Gestational History Variables* |  |  |  |  |
| Reason for genetic screening |  |  |  | 0.427 |
| Elective | 28 (39.4%) | 12 (34.3%) | 16 (44.4%) |  |
| Advanced Maternal Age | 41 (57.7%) | 22 (62.9%) | 19 (52.8%) |  |
| Abnormal US/Other | 2 (2.8%) | 1 (2.9) | 1 (2.9%) |  |
| Prenatal confirmatory testing | 37 (52.1%) | 16 (45.7%) | 21 (58.3%) | 0.408 |
| Maternal pre-pregnancy BMI (kg/m<sup>2</sup>) | 25.8 ± 5.5 | 25.2 ± 6.0 | 26.4 ± 5.0 | 0.376 |
| Maternal pregnancy weight gain (kg) | 13.1 ± 7.0 | 13.4 ± 7.2 | 12.8 ± 6.9 | 0.722 |
| Maternal age at birth (years) | 35.1 ± 5.1 | 34.9 ± 4.1 | 35.3 ± 5.9 | 0.713 |
| Paternal age at birth (years) | 35.9 ± 5.3 | 35.9 ± 5.1 | 36.0 ± 5.5 | 0.965 |
| Gestational age (weeks) | 39.3 ± 1.1 | 39.2 ± 1.0 | 39.4 ± 1.2 | 0.598 |
| Birthweight (kg) | 3.26 ± 0.42 | 3.24 ± 0.46 | 3.29 ± 0.39 | 0.613 |
| Birth length (cm) | 50.8 ± 2.5 | 50.6 ± 2.6 | 50.9 ± 2.4 | 0.625 |
| *Visit 1 Measures* |  |  |  |  |
| Infant age (days) | 65.5 ± 15.7 | 65.8 ± 16.0 | 65.1 ± 15.6 | 0.867 |
| Feeding source |  |  |  | 0.931 |
| Breast milk only | 38 (53.5%) | 18 (51.4%) | 20 (55.6%) |  |
| Formula only | 14 (19.7%) | 7 (20.0%) | 7 (19.4%) |  |
| Breast milk + formula | 19 (26.8%) | 10 (28.6%) | 9 (25.0%) |  |
| Parent-described temperament |  |  |  | 0.569 |
| Easy-Going | 42 (59.2%) | 21 (60%) | 21 (58.3%) |  |
| Average | 28 (39.4%) | 13 (37.1%) | 15 (41.7%) |  |
| Difficult | 1 (1.4%) | 1 (2.9%) | 0 (0.0%) |  |
| Weight z-score | -0.73 ± 0.92 | -0.72 ± 0.96 | -0.73 ± 0.88 | 0.962 |
| Length z-score | -0.59 (1.06) | -0.78 (0.94) | -0.41 (1.14) | 0.140 |
| Weight-for-length z-score | -0.26 (1.05) | -0.02 (1.05) | -0.50 (1.02) | 0.057 |
| Penile length (cm) | 3.71 (0.49) | 3.68 (0.45) | 3.73 (0.53) | 0.667 |
| Penile length z-score | -0.24 (0.61) | -0.28 (0.56) | -0.21 (0.67) | 0.667 |
| %fat mass z-score | -0.77 (1.04) | -0.77 (1.18) | -0.77 (0.90) | 0.999 |
| Fat free mass (grams) | 4.19 (0.55) | 4.22 (0.62) | 4.15 (0.47) | 0.626 |
| Fat mass (grams) | 1.05 (0.37) | 1.07 (0.42) | 1.04 (0.32) | 0.728 |
| Alberta Infant Motor Scale percentile | 31.4 (24.0) | 25.4 (23.9) | 37.2 (23.0) | 0.038 |
| Peabody gross motor quotient | 96.6 (5.7) | 96.3 (5.9) | 96.9 (5.6) | 0.689 |
| Peabody fine motor quotient | 91.6 (7.1) | 91.7 (7.4) | 91.5 (6.8) | 0.904 |
| Peabody total motor quotient | 93.8 (5.4) | 93.7 (5.4) | 93.9 (5.4) | 0.853 |
| Bayley 3 cognitive composite | 105 (9.9) | 106 (8.9) | 104 (10.8) | 0.280 |
| Bayley 3 language composite | 101 (8.0) | 102 (7.2) | 100 (8.7) | 0.238 |
| Bayley 3 motor composite | 101 (9.1) | 100 (8.3) | 101 (10.0) | 0.483 |
| Luteinizing Hormone (mIU/mL) | 3.74 [3.04, 4.86] | 3.71 [3.40, 4.51] | 4.04 [2.84, 5.00] | 0.531 |
| Follicle Stimulating Hormone (mIU/mL) | 2.05 [1.66, 2.88] | 1.89 [1.54, 2.43] | 2.30 [1.78, 2.89] | 0.136 |
| Total testosterone (ng/dL) | 175 [132, 207] | 173 [122, 207] | 178 [138, 200] | 0.724 |
| Inhibin B (pg/mL) | 248 [201, 301] | 242 [197, 300] | 256 [218, 301] | 0.878 |
| Anti-Mullerian Hormone (pmol/L) | 839 [634, 1107] | 877 [750, 1107] | 833 [625, 1107] | 0.564 |
| Data are represented as mean ± standard deviation; median [25<sup>th</sup> percentile, 75<sup>th</sup> percentile]; or n (%).<br>Group A received testosterone in the first half of the study and Group B received placebo.<br>SES = socioeconomic status | | | | |

The *a priori* primary outcome of change in %FM z-score at the 12-week visit was statistically different between the treatment groups, with the placebo-treated group gaining a %FM z-score of 0.56 more than the testosterone-treated group (Table 2). When examined by absolute %FM rather than z-score, this translated to a gain of 2.8 ± 4.7 percent in the testosterone-treated group compared to 4.8 ± 4.4 percent in the placebo group (p=0.081). Body composition differences were entirely due to FFM gain: +151 ± 43g in the testosterone-treated group versus +118 ± 35g in the placebo group (p=0.001); whereas FM gain did not statistically differ: +61 ± 36g vs +71 ± 36g (p=0.270). Testosterone treatment was associated with a robust linear growth velocity of 35.1 ± 6.9 cm/yr compared to 29.3 ± 5.9 cm/yr for infants receiving placebo (p<0.001). Stretched penile length increased by 1.1 ± 0.5cm (z-score +1.3 ± 0.6) with testosterone, compared to 0.1 ± 0.5cm (z-score +0.1 ± 0.6) in the placebo group (p<0.001).

**Table 2.** Change scores for primary and select secondary outcomes at the primary endpoint (12 weeks)

|  | **Testosterone** | **Placebo** | **Mean Difference<sup>^</sup>** | **p-value** |
| --- | --- | --- | --- | --- |
| %FM z-score | -0.07 ± 1.0<br>*n*=31 | 0.49 ± 1.0<br>*n*=30 | -0.56 [-1.1, -0.06] | 0.030* |
| Length z-score | 0.65 ± 0.67<br>*n*=34 | -0.02 ± 0.69<br>*n*=34 | +0.68 [0.35, 1.01] | <0.001* |
| Stretched penile length (cm) | 1.1 ± 0.5<br>*n*=34 | 0.1 ± 0.5<br>*n*=34 | +1.0 [0.78, 1.25] | <0.001* |
| AIMS %ile | 15.4 ± 30<br>*n*=35 | 8.4 ± 30<br>*n*=36 | +7.0 [-7.0, 21.0] | 0.319 |
| Bayley 3 Cognitive Composite | -3.4 ± 9.8<br>*n*=34 | -1.4 ± 13.5<br>*n*=34 | -2.0 [-7.8, 3.7] | 0.484 |
| Bayley 3 Language Composite | -7.0 ± 10.1<br>*n*=34 | -3.7 ± 9.7<br>*n*=33 | -3.4 [-8.1, 1.4] | 0.166 |
| Bayley 3 Motor Composite | -3.0 ± 14.0<br>*n*=34 | -6.0 ± 13.2<br>*n*=33 | +3.0 [-3.7, 9.6] | 0.375 |
| PDMS-2 Gross Motor Quotient | 1.8 ± 7.4<br>*n*=34 | 0.1 ± 7.2<br>*n*=33 | +1.7 [-1.8, 5.3] | 0.335 |
| PDMS-2 Fine Motor Quotient | 9.0 ± 9.5<br>*n*=32 | 6.7 ± 9.3<br>*n*=34 | +2.3 [-2.3, 6.9] | 0.325 |
| ABAS-3 Generalized Adaptive Composite | 3.5 ± 8.1<br>*n*=35 | 2.9 ± 7.4<br>*n*=36 | +0.6 [-3.1, 4.3] | 0.747 |
| Data are represented as mean ± standard deviation. <sup>^</sup>Absolute Mean Difference and 95% confidence interval of the mean difference between testosterone-treated vs placebo-treated groups. FM = fat mass; AIMS = Alberta Infant Motor Scales; PDMS-2 = Peabody Developmental Motor Scales - Version 2 (quotients ; ABAS-3 = Adaptive Behavior Assessment System Third Edition<br>*denotes significance at alpha of 0.05 |  |  |  |  |

In contrast to these physical outcomes, the *a priori* outcome of AIMS total score assessing gross motor abilities was not different between treatment groups (change in AIMS percentile score 15.4 ± 29.5 vs 8.4 ± 29.6, p=0.319). Cognitive, language, and motor composites and their respective subdomains on the Bayley-3 were not different between treatment groups at Visit 2 (Table 2 and Figure 2). Motor outcomes assessed by the PDMS-2 were also not different between treatment groups. There were also no differences in scores for any of the domains on parent-rated adaptive function assessment (ABAS-3).

**FIGURE 2.**
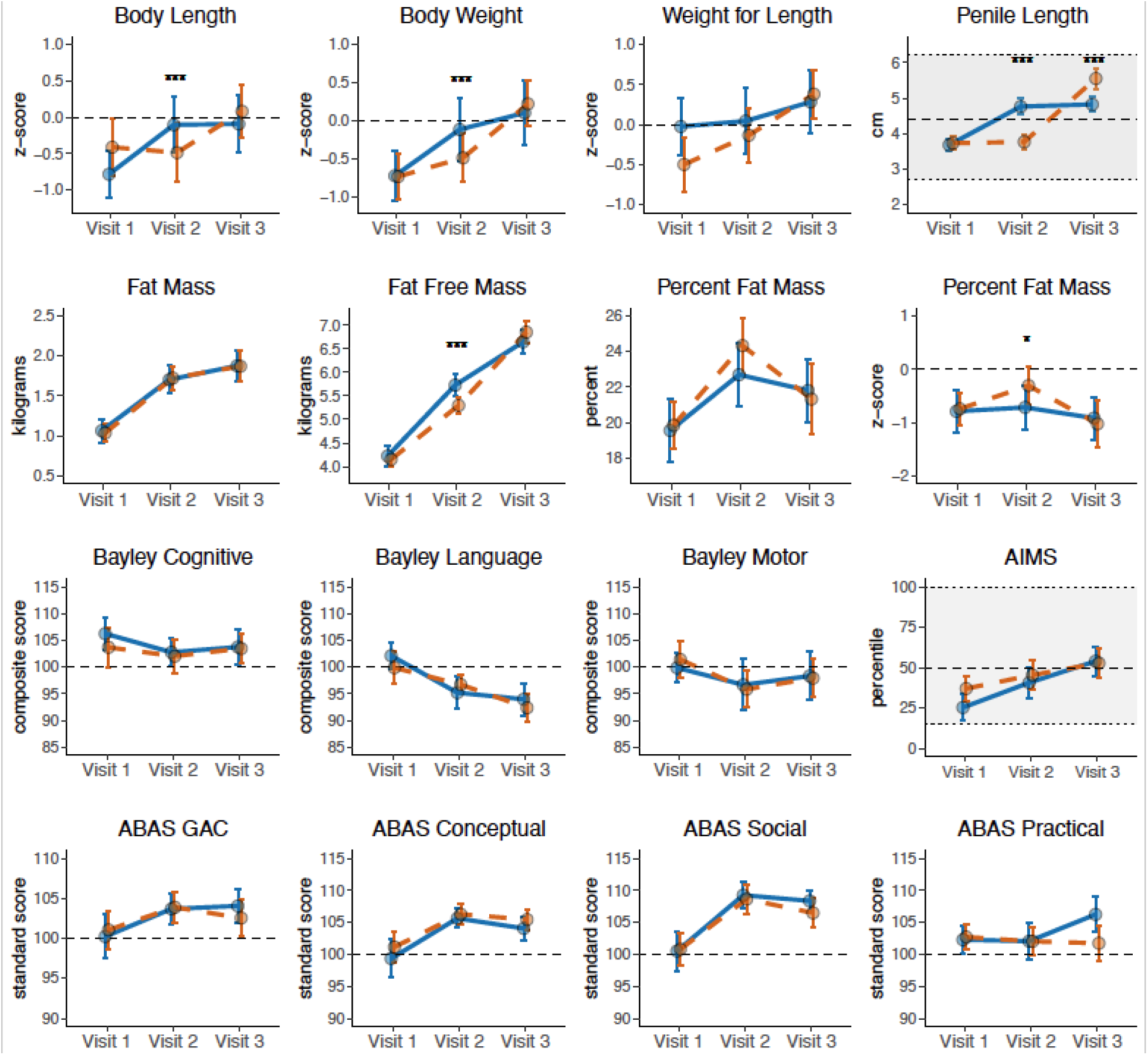
Mean and 95% confidence interval of the mean by study visit for Group A (blue line; testosterone first, placebo second) and Group B (orange dashed line, placebo first, testosterone second) at Visit 1 (baseline, ∼2 months of age), Visit 2 (∼5 months of age), and Visit 3 (∼8 months of age). Outcomes from blinded physical examination (row 1), PEAPOD air displacement plethysmography (row 2), neurodevelopmental assessment (row 3), and parent-reported development (row 4). Where applicable, dashed line represents the average in the general population and normal ranges are depicted with a shaded background. Significant differences between groups is indicated by asterices (* = <0.05, ** = <0.01, *** = <0.001). AIMS = Alberta Infant Motor Scales; ABAS = Adaptive Behavior Assessment System Third Edition

Testosterone treatment suppressed reproductive hormone concentrations (Figure 3). The change in LH between visit 1 and 2 for group A was -3.5 mIU/mL [-4.1, -2.9] vs -1.8 [-3.3, -0.8] in group B, p<0.001). FSH (-1.5 mIU/mL [-2.0, -1.0] vs -0.7 [-1.3, -0.4], p<0.001) and INHB (-113 pg/mL [-167, -66] vs 0.0 [-31.7, 31.0], p<0.001) were also significantly suppressed with testosterone treatment. There was not a significant difference between groups for change in AMH (+128 pmol/L [0, 401] vs +174 [-0.5, 312], p=0.669).

**Figure 3.**
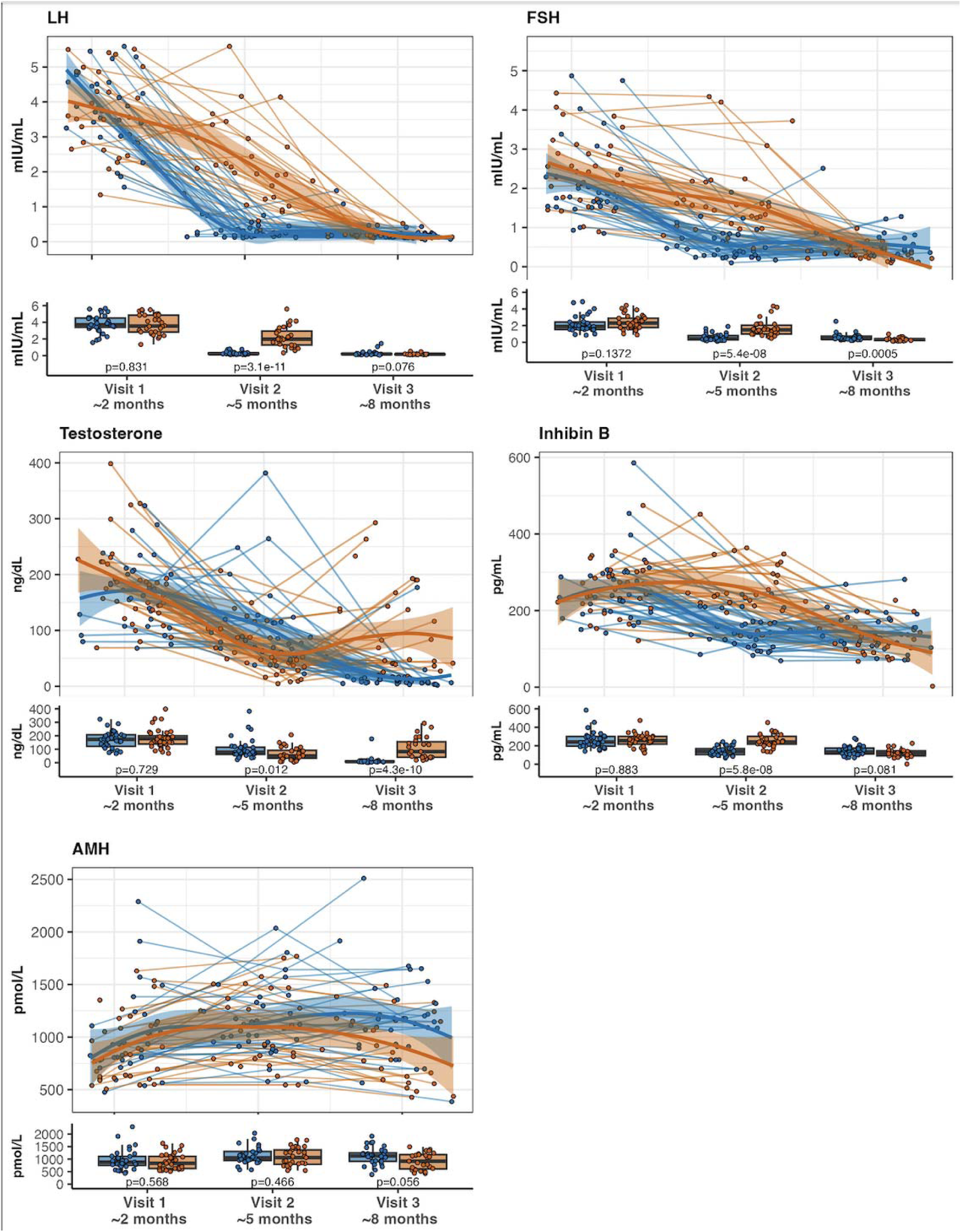
Spaghetti plots for longitudinal hormone concentrations by age (1-9 months) and corresponding box plots at each study visit for those in group A (blue) treated with testosterone followed by placebo, and group B (orange) treated with placebo followed by testosterone. Each circle is a value for an individual participant at a given time; lines of the spaghetti plot represent individual participants over time, and the bolder lines are smoothed loess curves with 95% confidence intervals around the loess curves for each group. Box plots show the 1^st^ and 3^rd^ quartiles as the bottom and top of the box respectively with median in the middle and error bars representing 95% of the data. LH = luteinizing hormone, FSH = follicle stimulating hormone, AMH = anti-mullerian hormone.

Results for the second half of the study (changes from Visit 2 to Visit 3, blinded treatment cross-over), were similar to the first half of the study. Testosterone treatment increased FFM and both body and penile length with no discernable effect on any neurodevelopmental outcomes. At Visit 3, the group receiving testosterone in the second half of the study had a significantly greater penile length (5.6 ± 8.4 cm vs 4.8 ± 6.0 cm, p<0.001), lower FSH (0.32 [0.21, 0.41] vs 0.44 mIU/mL [0.37, 0.73], p<0.001), and borderline significantly lower AMH (918.00 [609.50, 1178.00] vs 1145.00 [910.00, 1319.00], p=0.038; there were no differences in any other outcomes. When evaluating change scores while on testosterone, Testosterone treatment was not associated with a difference in parent-reported temperament. At Visits 2 and 3 respectively, 86% and 82% of infants were rated by their parent(s) as “easy-going” with the remainder being “average” temperament (none rated as “difficult”), with no differences based on testosterone treatment.

Results did not change when regression models were applied controlling for potential confounding variables (e.g. race, ethnicity, SES, breastfed status, maternal BMI or pregnancy weight gain, birthweight). Similarly, neither baseline testosterone concentration nor baseline inhibin B concentrations influenced the relationship between testosterone treatment and any developmental outcomes.

During the 6-month study period all infants experienced at least one adverse event for a total of 483 adverse events reported, with 297 occurring during testosterone administration and 210 during placebo administration (Table 3). Increased penile erections was the only adverse event occurring in significantly more individuals while on testosterone (62.9% of participants on testosterone vs 11.3% participants on placebo, p<0.001). However, pubic hair (14.1% of participants) and acne (47.9% of participants) were also attributed to testosterone treatment for biologic plausibility. During testosterone administration, two patients had emergency room encounters unrelated to the study, one surgery unrelated to the study, no hospitalizations and no deaths. During placebo administration, there were a total of seven emergency room encounters unrelated to the study and two surgeries unrelated to the study, no hospitalizations and no deaths.

**Table 3.** Adverse events.

| Adverse Events (AE) | Entire Study |  | On Testosterone |  | On Placebo |  | RR | 95% CI | P value |
| --- | --- | --- | --- | --- | --- | --- | --- | --- | --- |
|  | # of AEs | Indiv (%) | # of AEs | Indiv (%) | # of AEs | Indiv (%) |  |  |  |
|  | n = 488 | n = 71 | n = 278 | n = 70 | n = 210 | n = 71 |  |  |  |
| Any AE | -- | 68 (95.8%) | -- | 64 (91.4%) | -- | 56 (80.0%) | 1.87 | 1.03 - 3.93 | 0.057 |
| Increased Fussiness | 91 | 45 (63.4%) | 54 | 34 (48.5%) | 37 | 26 (36.6%) | 1.28 | 0.91 - 1.77 | 0.175 |
| **Penile Erections** | 87 | 44 (62.0%) | 71 | 44 (62.9%) | 16 | 8 (11.3%) | 2.90 | 2.08 - 4.14 | **<0.001** |
| Increased Appetite | 88 | 39 (54.9%) | 43 | 27 (38.6%) | 45 | 30 (42.3%) | 0.93 | 0.65 - 1.29 | 0.732 |
| **Acne or boils** | 62 | 34 (47.9%) | 33 | 23 (32.9%) | 29 | 20 (28.2%) | 1.12 | 0.77 - 1.55 | 0.586 |
| Increased Sleepiness | 54 | 30 (42.3%) | 28 | 19 (27.1%) | 26 | 18 (25.4%) | 1.05 | 0.70 - 1.47 | 0.850 |
| Eczema or another rash | 30 | 26 (36.6%) | 13 | 12 (17.1%) | 17 | 16 (22.5%) | 0.84 | 0.50 - 1.26 | 0.527 |
| Fever | 16 | 11 (15.5%) | 6 | 5 (7.1%) | 10 | 8 (11.3%) | 0.76 | 0.34 - 1.32 | 0.562 |
| Injection site reaction | 13 | 9 (12.7%) | 7 | 7 (10%) | 6 | 3 (4.2%) | 1.46 | 0.81 - 2.02 | 0.208 |
| Vomiting | 12 | 9 (12.7%) | 5 | 5 (7.1%) | 7 | 6 (8.5%) | 0.91 | 0.42 - 1.51 | 0.999 |
| **Pubic hair** | 10 | 10 (14.1%) | 6 | 6 (8.6%) | 4 | 4 (5.6%) | 1.23 | 0.63 - 1.82 | 0.532 |
| Change in stool pattern | 7 | 6 (8.5%) | 5 | 4 (5.7%) | 2 | 2 (2.8%) | 1.36 | 0.61 - 1.99 | 0.441 |
| Upper respiratory infection | 7 | 6 (8.5%) | 2 | 2 (2.9%) | 5 | 5 (7.0%) | 0.56 | 0.16 - 1.30 | 0.441 |
| Acute otitis media | 4 | 4 (5.6%) | 1 | 1 (1.4%) | 3 | 3 (4.2%) | 0.50 | 0.09 - 1.43 | 0.620 |
| Sleeplessness | 4 | 2 (2.8%) | 3 | 1 (1.4%) | 1 | 1 (1.4%) | 1.01 | 0.19 - 1.92 | 0.999 |
| Decreased Appetite | 2 | 2 (2.8%) | 0 | 0 (0%) | 2 | 2 (2.8%) | 0.00 | 0.00 - 1.33 | 0.497 |
| Edema of Foreskin | 1 | 1 (1.4%) | 1 | 1 (1.4%) | 0 | 0 (0%) | 2.03 | 0.42 - 9.60 | 0.497 |

AE = adverse event; Indiv = individual participants; RR = risk ratio for adverse event occurring while on testosterone compared to while on placebo. AEs could be reported at any time throughout the 6-month study period. Bolded AEs were attributed to testosterone treatment.

## DISCUSSION

In this randomized, double-blind, placebo-controlled clinical trial, we found that three doses of testosterone cypionate 25 mg given intramuscularly every four weeks to infants with 47,XXY induce anabolic changes in growth and body composition. These physical outcomes were overall favorable, as they normalized penile length, growth velocity, and lean mass in the short term. However, this intervention did not yield measurable beneficial effects on neurodevelopment, even in post hoc subgroup analyses. Additionally, our findings suggest potential adverse effects on the endogenous HPG axis, challenging the assumption that a short course of testosterone treatment is benign. Based on these results, there is no evidence on which to recommend testosterone treatment to modify the neurodevelopmental trajectory for infants with XXY; however, we cannot exclude a possible effect on neurodevelopmental outcomes emerging over time given the short evaluation time in this study. Longer term evaluation of cardiometabolic, neurodevelopment, and gonadal function outcomes is needed.

The physical effects of testosterone treatment that we observed align with our pilot study that found an increase in lean mass and corresponding lower adiposity.(29) Multiple observational studies have found higher adiposity in XXY compared to male controls throughout the lifespan.(29,39,40) In addition, adiposity early in life is associated with cardiometabolic disorders in adulthood, potentially due to lower lean mass, greater fat mass, or both.(41,42) The favorable results we observed on body composition may or may not persist and/or impact later metabolic health in this cohort, and require further study. Similarly, testosterone treatment significantly increased penile size. A short course of testosterone is currently used in clinical practice to treat micropenis, resulting in increase in penile length with few reported side effects.(43) Upon enrollment in the current study, we found penile length to be slightly shorter than average, however none of the boys enrolled in this study would qualify as having micropenis. While treatment of micropenis with testosterone is accepted in both endocrinology and urology clinical practice, there are limited data informing short- or long-term outcomes of micropenis treatment beyond the initial gain in penile size, including the impact on future testicular function, childhood/adult wellbeing, or adult penile length. These longer-term outcomes are likely of more importance to patients and parents and warrant future study.

Our results are in contrast to existing literature proposing that testosterone treatment in the first year of life is associated with better neurodevelopmental outcomes in childhood in XXY. Despite assessing multiple developmental domains with several standardized assessments and parent report, we did not appreciate any evidence supportive of neurodevelopmental benefits, despite multiple *post-hoc* subanalyses. Samango-Sprouse et al. compared a cohort of boys with XXY seen for clinical developmental evaluation who received infant testosterone with those who did not, reporting cognition, motor ability, language development, social skills, and behavior are all superior in the group with infant testosterone exposure.(19,44–46) However, there are multiple confounding factors that may have had an impact on these findings, including but not limited to baseline differences in families seeking off-label treatments (no randomization) and the lack of blinding of the assessors of subjective outcomes. Pursuing infant testosterone may have a positive effect on parents (e.g. hope, empowerment) and subsequently parent-child attachment that supports development – we did not assess these outcomes. It is also possible that the many neurodevelopmental benefits found by Samango-Sprouse et al are not immediately apparent but emerge with time. Further study will be needed to answer these questions.

Treatment emergent adverse events were generally anticipated, minor, and temporary, with the exception of suppression of the endogenous HPG axis that may have long-term effects on testicular function and future spermatogenesis. While testicular dysfunction and infertility are nearly universal in XXY, most will spontaneously enter puberty and up to half of young men seeking biologic paternity can successfully retrieve sperm through testicular sperm extraction. Evaluation of testicular function in childhood, puberty, and beyond will be needed before we can consider infant testosterone a safe intervention for XXY or other indications. Testosterone treatment did not negatively affect infant temperament, which continued to rated by parents as “easy going”.

While this study was robustly designed and adequately powered for short-term outcomes, there are several limitations to consider before applying these findings to practice. First, although we achieved diversity in terms of self-reported race and ethnicity, this study sample was highly educated and socioeconomically affluent, not only affecting generalizability to other populations but also potentially minimizing variability in outcomes that may mask smaller yet potentially significant benefits from testosterone. Next, the neurodevelopmental outcome measures used in this study may not be sensitive enough to detect changes in this population, although our inclusion of three different standardized direct assessments and a parent-report measure strengthens our confidence in our null findings for neurodevelopmental outcomes. Given we had very few boys with low baseline testosterone concentrations, it is still plausible those with an insufficient testosterone surge during the mini-puberty period would have benefits from supplemental testosterone. Finally, this study was designed to assess short-term outcomes only and these outcomes do not necessarily have immediate clinical implications. Longitudinal follow up will be needed to determine the duration of the observed physical and hormonal effects of testosterone treatment in infancy, as well as establish whether neurodevelopmental benefits emerge over time.

## Conclusion

In conclusion, in this double-blind RCT, testosterone treatment in infants with XXY had clear effects on physical outcomes and was well-tolerated clinically, however no benefits to short-term neurodevelopmental outcomes were noted. Testosterone suppressed the HPG axis including the production of inhibin B, with unknown implications for future testicular function. The results of this study do not support universally adopting testosterone treatment for infants with XXY into clinical practice at this time. Long term follow up is needed to determine if the altered hormone profile persists, if the advantageous effects on body composition are sustained, and if neurodevelopmental benefits emerge with time or in a specific subset of the population.

## Statement of Ethics

This study protocol was reviewed and approved by the Colorado Multiple Institutional Review Board, #17-1317. Written informed consent was obtained by a parent or legal guardian prior to any study procedures. The study was registered on clinicaltrials.gov (NCT03325647).

## Data availability

Deidentified individual-level data are deposited in the NIH Data and Specimen Hub (DASH), a controlled-access data repository, and can be requested through the DASH Portal (dash.nichd.nih.gov).

## ACKNOWLEDGEMENTS

First and foremost, the study team thanks the families who participated in this research, many of whom had to travel to the study site with their newborns during the COVID-19 pandemic. We also acknowledge the clinicians who referred their patients to the study, our patients who inspired this research, and all our colleagues in the eXtraOrdinarY Kids Clinic and Research Program. Thank you to Mariah Brown and Amira Herstic for study coordination. This study was supported by the National Institute of Child Health and Human Development (NICHD K23HD092588) and the (NIH/NCATS Colorado CTSA Grant Number UM1 TR004399 and the Department of Pediatrics at the University of Colorado Anschutz Medical Campus. The contact is solely the responsibility of the authors and does not necessarily represent the official views of the National Institutes of Health.

