## Supplemental methods for "Testosterone Effects on Short-Term Physical, Hormonal, and Neurodevelopmental Outcomes in Infants with 47,XXY/Klinefelter Syndrome: The TESTO Randomized Controlled Trial"

**Supplemental Materials: Hormone measurement methods**

Testosterone was measured using liquid chromatography-tandem mass spectrometry (LC-MS/MS). One hundred µl of serum (sample or QC control) were vortex-mixed with 20 µl of the internal standard solution (testosterone -1a,2a-D2) and 1.5 ml ethyl ether for 3 min. The solutions were then allowed to stand for 20 min, when the supernatants were separated and evaporated to dryness at room temperature under a stream of nitrogen. The dried extracts were reconstituted in 100 µl MeOH/water (50/50; V/V). The samples were injected into an Acquity® UPLC system (Waters, Manchester, UK), equipped with an Acquity® UPLC BEH C18 column (Waters, Guyancourt, France). The mobile phase consisted of a mixture of water and MeOH. It was operated with a flow-rate of 0.3 ml/min in gradient mode, at a temperature of 40°C. Mass spectra were recorded using a Xevo TQ-XS® triple quadrupole mass spectrometer (Waters, Manchester, UK). Measurements were performed using positive electrospray ionization (ESI) in Multiple Reaction Monitoring (MRM) data-acquisition mode. The parameters of the electrospray interface were optimized as follows: capillary voltage 3 kV, source temperature 150°C, desolvatation temperature 650°C, desolvatation gas 1000 l/hr.

Steroid identification was based on an identical retention time and two identical mass transitions with authentic reference compound (Testosterone ChromSystems, ref 72040). Quantification was performed relative to a calibration series of 7 points (ref cal 3523: 1.9, 4.5, 26.5, 99.2, 292, 577, 1160 ng/dL). Data processing was ensured using MassLynx software (V 4.2, SCN 982, Waters, Manchester, UK). This software was used to detect and integrate the peaks. Accuracy checked in our unit did not differ from 100%. Detection limit was 0.02 nmol/l. Intra-assay coefficients of variation (CV) were <6% at detection level, and <5% throughout the range of observed values. Inter-assay CVs were <8 % throughout the range of observed values. Internal quality controls (Chromsystems, ref 0338, 0339, 0340) at 3 different levels, low 21.8, medium 149 and high 770 ng/dL, are run in each series at 3 different positions: at the top, the middle and the end of the series. Moreover, 6 times in the year, external quality controls, provided by ProBioQual, Lyon, France, were additionally run in a blind way in the Testosterone series and their results peer-reviewed, thanks to a voluntary subscription to this national quality control system.

Serum levels of Inhibin B were measured by means of a solid-phase sandwich assay using Ansh Labs reagents (Webster, TX) (Ansh Labs cat. n° AL-107, RRID:AB_2783661; https://www.antibodyregistry.org/AB_2783661). Inhibin A exhibited 1% cross-reactivity in the inhibin B assay. The quantification range was 4 to 1100 pg/ml. The measurements were extrapolated from the calibration dose-response data in a non-linear method using a cubic spline curve. The intra- and inter-assay CVs were 6.8 and 10.5 % respectively at the level of 39 pg/mL, and 5.7 and 8 % respectively at the level of 112 pg/mL. The detection limit was 4.6 pg/mL. Internal quality controls, one low at approximately 100 and one high, at approximately 400 pg/ml, provided by Ansh Labs, are run in each series. Additionally, in each series we run a pool of post-menopausal women, previously checked as being free from Inhibin B, to assess the absence of any false positive signal. Moreover, 6 times in the year, external quality controls, provided by ProBioQual, Lyon, France, were additionally run in a blind way in the series and their results peer-reviewed.

AMH levels were measured by means of a solid-phase sandwich assay using Ansh Labs reagents (Webster, TX) (Ansh Labs cat. n° AL-105, RRID:AB_2783659; https://www.antibodyregistry.org/AB_2783659). There was no cross-reactivity of related proteins including Transforming Growth Factor beta. The quantification range was: 0.50 to 114 pmol/L. The measurements were extrapolated from the calibration dose-response data in a non-linear method using a cubic spline curve. The intra- and inter-assay CVs were 2.3 and 3.1% respectively at the level of 107 pmol/ L, and 1,4 and 2,5% respectively at the level of 557 pmol/L. The detection limit was 0.7 pmol/L. Internal quality controls, one low and one high, provided by Ansh Labs, are run in each series. Additionally, in each series we run a pool of post-menopausal women, previously checked as being free from AMH, to assess the absence of any false positive signal. Moreover, 6 times in the year, external quality controls, provided by ProBioQual, Lyon, France, were additionally run in a blind way in the series and their results peer-reviewed.

FSH and LH were measured by means of a sensitive electro-chemiluminescent immunoassay (ECLIA) intended for use on a Cobas 6000 immuno-analyzer (Roche Diagnostics, Meylan, France). The ECLIA Kit for FSH from Cobas Elecsys contains: Biotinylated monoclonal anti-FSH antibody (mouse) and Monoclonal anti-FSH antibody (mouse) labeled with ruthenium complex (RRID:AB_2800499). In this FSH assay, the cross-reactivity of related proteins LH, TSH, hCG, hGH, hPL was < 0.1 %. The quantification range was 0.1 to 200 mIU/mL. Two internal quality controls at 2 different levels (Precicontrol Universel ref 11731416190 from Roche Diagnostics) were run in each series to ensure stability of the specimens and to determine imprecision. Moreover, 6 times in the year, external quality controls, provided by ProBioQual, Lyon, France, were additionally run in a blind way in the series and their results peer-reviewed. The intra- and inter-assay CVs were respectively 2.01 and 3.15 % at the level of 2.95 mIU/mL

The ECLIA Kit for LH from Cobas Elecsys contains: Biotinylated monoclonal anti-LH antibody (mouse) and Monoclonal anti-LH antibody (mouse) labeled with ruthenium complex (RRID:AB_2800498). In this LH assay, the cross-reactivity of related proteins FSH, TSH, hCG, hGH, hPL was < 0.1 %. The quantitation range was 0.1 to 200 mIU/mL. Two internal quality controls at 2 different levels (Precicontrol Universel ref 11731416190 from Roche Diagnostics) were run in each series to ensure stability of the specimens and to determine imprecision. The intra- and inter-assay CVs were respectively 1.94 % and 3.08 % at the level of 2.95 mIU/mL. The detection limit was 0.1 mIU/mL for both assays.
